# Cost and cost-effectiveness of swab-based molecular testing for tuberculosis in the Philippines, Uganda, Vietnam, and Zambia

**DOI:** 10.1101/2025.08.14.25333714

**Authors:** Armen Jheannie D. Barrameda, Penelope Papadopoulou, Charles Yu, Ha Phan, Monde Muyoyeta, Abigail K. de Villiers, Hien Le, Seke Muzazu, Pia A. Steimer, Seda Yerlikaya, Adithya Cattamanchi, Claudia M. Denkinger, Alfred Andama, Florian M. Marx

## Abstract

**Background:** Tongue swabs (TS) offer a promising alternative to sputum-based molecular testing for tuberculosis. As part of the Tongue Swab Yield (TSwaY) study, we assessed the cost and cost-effectiveness of integrating TS-based testing using MiniDock MTB (Guangzhou Pluslife Biotech Co., Ltd., China) into primary healthcare in four high-burden countries.

**Methods:** Cost data were collected from primary healthcare facilities in the Philippines, Uganda, Vietnam, and Zambia. We evaluated three MiniDock MTB strategies: (1) *TS-only*, replacing sputum Xpert Ultra; (2) *limited combined*, with Xpert Ultra first-line and TS added for sputum-scarce individuals; (3) *extended combined*, with TS also added for those with negative or indeterminate sputum results. Additionally, we simulated an *integrated combined* strategy using MiniDock MTB to test sputum swabs (if available) and TS for sputum-scarce individuals. Incremental cost-effectiveness ratios (ICERs) for each scenario were estimated relative to the next least costly strategy. Net monetary benefits (NMB) were evaluated across willingness-to-pay (WTP) thresholds. Best estimates were based on observed diagnostic yield, with 95% uncertainty intervals from probabilistic simulation.

**Findings:** Among 1370 participants, *TS-only* was least costly but yielded the fewest diagnoses (62 [95% UI: 48;75] at USD 300 [95% UI: 244;388] per diagnosis). The standard of care, *sputum Xpert Ultra*, was extendedly dominated by TS-based strategies. *Limited combined* yielded 12 (95% UI: -1;22) additional diagnoses at an incremental USD 1,507 per diagnosis compared with TS-only; *extended combined* yielded 15 (95% UI: 8;22) additional diagnoses at an incremental USD 1,004 (95% UI: 661;1,655) per diagnosis compared with limited combined. The simulated *integrated combined* sputum swab and TS testing using MiniDock MTB had the highest NMB at WTP thresholds above USD 161 per additional diagnosis.

**Interpretation:** Our findings support integrating swab-based testing with MiniDock MTB into diagnostic algorithms in high-burden settings to increase case detection at minimal additional cost.

**Funding:** Gates Foundation.

## Introduction

Rapid molecular tests offer substantial advantages over traditional diagnostic methods for the bacteriological confirmation of tuberculosis (TB), raising hopes of improving case detection and accelerating access to curative treatment. Scaling up molecular testing has therefore been a key priority for national TB programmes over the past decade. Widespread adoption of Xpert MTB/RIF and its successor, Xpert MTB/RIF Ultra (“Xpert Ultra”; Cepheid, USA), has enhanced TB detection in routine settings.^1,2^ However, high costs, infrastructure limitations, and poor adherence to diagnostic algorithms have constrained implementation and impact.^1-3^

A key challenge for currently recommended rapid molecular tests is their reliance on sputum samples. In high TB burden countries, several studies have reported sputum scarcity, defined as the inability to expectorate sputum for diagnostic evaluation, in 18–35% of adults evaluated for TB.^4-7^ These limitations contribute to the suboptimal diagnostic yield of sputum-based diagnostics and highlight the urgent need for accurate and affordable non-sputum diagnostic tools to support earlier detection and treatment.^8,9^

Tongue swabs have emerged as a promising alternative to sputum-based molecular testing for TB, particularly because they are easier and more acceptable to collect.^10-13^ Building on this, novel swab-based near–point-of-care (POC) platforms have been developed.^14^ One such system is the MTB Nucleic Acid Test Card (MiniDock MTB; Guangzhou Pluslife Biotech, China), a qualitative molecular assay designed to detect the *Mycobacterium tuberculosis* complex (MTBC). The test runs on the Pluslife Integrated Nucleic Acid Testing Device (MiniDock PM001 Ultra), a compact, isothermal platform that enables rapid, low-cost testing. Sample preparation is handled by the Pluslife Thermolyse, an automated unit combining bead-beating and thermal lysis to directly process sputum or tongue swab specimens.^14,15^ Early diagnostic accuracy studies have shown that tongue swab assays have high specificity and sensitivity exceeding that of sputum smear microscopy, though lower than sputum Xpert Ultra.^14^ However, tongue swab-based testing could improve diagnostic yield when offered to individuals who are unable to produce sputum and would otherwise remain untested with sputum-based assays.^7,8^

Beyond test performance, economic considerations are essential to guide implementation decisions.^16^ MiniDock MTB is anticipated to be substantially less expensive than current World Health Organization (WHO)-recommended sputum-based diagnostics, offering potential cost savings through lower device and cartridge costs, reduced staffing needs, and simpler logistics.^14^ Its operational simplicity may also support decentralized implementation, particularly in peripheral health settings. As a result, tongue swab-based molecular testing using MiniDock MTB could represent a more affordable and accessible alternative to sputum-based approaches. However, empirical evidence on the cost and cost-effectiveness of this approach remains limited. Early health-economic evaluations are needed to inform optimal integration into routine TB diagnostic services.

This health-economic evaluation was conducted as part of the Tongue Swab Yield (TSwaY) study, a multi-country, pragmatic cross-sectional study in primary healthcare centers that assessed novel tongue swab-based approaches using the MiniDock MTB platform.^17^ We aimed to evaluate the cost and cost-effectiveness of integrating tongue swab-based testing using MiniDock MTB into primary healthcare in four high-burden countries. In addition, we used simulation to assess the health and cost implications of an integrated, single-platform diagnostic strategy in which MiniDock MTB would be used to test sputum swabs from individuals able to produce sputum and tongue swabs from those who were sputum-scarce.

## Methods

### Study Overview and Design

We conducted a health-economic evaluation of tongue swab-based molecular TB testing using MiniDock MTB in four high-burden countries. Details of TSwaY study procedures have been reported elsewhere.^17^ We collected cost data to compare tongue swab MiniDock MTB with sputum Xpert Ultra and assessed the cost-effectiveness of integrating tongue swab-based testing into routine diagnostic services. We followed the Consolidated Health Economic Evaluation Reporting Standards to report the results of our study.^18^

### Study population and setting

The TSwaY study was conducted in outpatient health centers providing primary diagnostic and treatment services for individuals with presumptive TB in the Philippines, Vietnam, Uganda, and Zambia. A total of 1,639 individuals who presented to study health centers were enrolled based on either unexplained cough for two or more weeks, or a TB risk factor (TB contact, history of mining, or HIV infection), plus an abnormal WHO-endorsed TB screening test (chest x-ray, or for PLHIV, serum C-reactive protein [CRP] >5 mg/dL).^17^ Individuals who were presenting for TB treatment after being diagnosed with TB elsewhere, already received treatment for TB, started on treatment for extra-pulmonary TB only, or were unable or unwilling to provide informed consent were excluded.^17^ For this cost-effectiveness analysis, we included only participants aged 15 years and older (n = 1,370), representing 84% of the total study population. This age restriction was applied as diagnostic procedures and performance differ in children, and the study was not powered for this subgroup.

### Interventions and comparator

This analysis compared three strategies for integrating tongue swab-based testing into routine diagnostic services for the bacteriological confirmation of TB, using observed yield data from the TSwaY study (*Supplementary Table S1*).

First, we evaluated a *tongue swab-only* strategy, in which sputum Xpert Ultra would be fully replaced by tongue swab testing using MiniDock MTB. Because MiniDock MTB does not detect rifampicin resistance, we assumed a reflex testing algorithm: individuals with a positive tongue swab result would undergo sputum Xpert Ultra testing for rifampicin resistance, where possible. For those unable to provide sputum, no further drug resistance testing was assumed.

Second, under the *limited combined* strategy, sputum Xpert Ultra would be used for individuals able to produce sputum, while tongue swab MiniDock MTB would be offered only to those unable to do so.

Third, under the *extended combined* strategy, both sputum (where possible) and tongue swab samples would be collected from all individuals. Sputum would be tested using Xpert Ultra, and tongue swab MiniDock MTB would be used for individuals unable to produce sputum (as in the limited strategy) and for those with negative or indeterminate sputum results.

All three strategies were applied retrospectively to individual-level data from the TSwaY study to compare diagnostic yield and cost with those of sputum Xpert Ultra alone, the current *standard of care*. Consistent with the TSwaY study protocol, only the test result of each participant’s first sample was included, and trace-positive Xpert Ultra results were excluded from the definition of bacteriologically confirmed TB. All testing was assumed to be conducted onsite.

### Simulated integrated sputum swab and tongue swab strategy

In a fourth, simulated scenario, we evaluated an integrated single-platform diagnostic approach in which MiniDock MTB would be used to test sputum swabs as the primary sample type, with tongue swabs collected from individuals unable to produce sputum. This “*integrated combined*” strategy was included to explore the health and cost implications of a same-platform approach that does not rely on Xpert Ultra. Operationally, sputum would be requested from all individuals, and a swab dipped into the sputum specimen would be tested using MiniDock MTB. Those unable to produce sputum would instead provide a tongue swab for MiniDock MTB testing. This strategy closely mirrors the *limited combined* strategy but replaces sputum Xpert Ultra with sputum swab MiniDock MTB testing.

Per-test cost estimates for sputum-swab MiniDock MTB testing were derived from the primary costing analysis. Because sputum swab testing was not conducted in the TSwaY study, diagnostic yield was estimated post hoc by applying an estimate of relative yield compared with sputum Xpert Ultra. This relative yield was sampled from pooled data from two early evaluations of sputum-swab MiniDock MTB accuracy conducted within the R2D2 TB Network (www.r2d2tbnetwork.org). One of these evaluations has been published,^19^ and the other is documented in an unpublished internal report shared confidentially with the study team. In these evaluations, sputum swab MiniDock MTB results were concordant with sputum Xpert Ultra results in 62 of 64 and 285 of 304 participants, respectively. Further methodological details are provided in the *Supplementary Information*.

### Costing and cost-effectiveness analysis

We estimated the cost of sputum Xpert Ultra and tongue swab MiniDock MTB-based molecular testing per individual evaluated for TB, expressed in 2024 US dollars, from a healthcare system perspective. Average per-test costs reflected 2024 estimates for TB diagnostic services in each of the four study countries and included human resources, consumables for sample collection and processing, device purchase and maintenance, and test cartridges. Salary and time inputs were derived from site data and staff interviews. All costs were converted to 2024 US dollars using average 2024 exchange rates from the International Monetary Fund; no inflation adjustment was required.^20^

Probabilistic per-test cost estimates were derived by sampling from specified ranges of itemized cost inputs. Using diagnostic yield and sample provision data from the TSwaY study, we calculated total costs for each strategy.

We first assessed the three diagnostic strategies directly informed by empirical yield data from TSwaY. We then extended the analysis to include the simulated *integrated combined* strategy as an alternative to the limited combined strategy. The *extended combined* strategy was not included in this comparison because no data were available to estimate the diagnostic yield of sequential testing with MiniDock MTB using both sputum and tongue swabs among people with negative and indeterminate sputum results. This precluded a meaningful extension of the *extended combined* strategy to a single-platform scenario.

The primary outcome was the incremental cost-effectiveness ratio (ICER), defined as the incremental cost per additional bacteriologically confirmed TB diagnosis, compared with the next least costly, non-dominated strategy. Best estimates reported were based on observed diagnostic yield, or simulated yield for the integrated combined strategy. Uncertainty intervals were generated using Monte Carlo simulation with 1,000 probabilistic model iterations. For each iteration, TB diagnoses were resampled using a nonparametric paired bootstrap at the individual level, preserving within-person correlation between sputum and tongue swab results.^21^ Sampled diagnostic outcomes were then combined with independently sampled cost estimates. We report 95% uncertainty intervals (95% UI) as the 2.5th and 97.5th percentiles of the resulting distribution.

We also estimated the probability that each strategy would be optimal at different willingness-to-pay (WTP) threshold per bacteriologically confirmed TB diagnosis. A strategy was defined as optimal at WTP threshold ω if it maximized net monetary benefit (NMB), calculated as ω × (number of diagnoses) – (total cost). The probability of each strategy being optimal was derived from the Monte Carlo simulation.

Analyses were conducted for the overall study population aged ≥ 15 years, as well as separately for each country.

In a sensitivity analysis, we accounted for incomplete sputum processing – particularly in Zambia – by assuming that unprocessed sputum samples had the same positivity rate as processed ones and recalculated diagnostic yield and ICERs accordingly.

### Ethical considerations

Written informed consent was obtained from all study participants in the TSwaY study. Ethical approval, including for the health-economic analysis, was obtained from institutional review boards and/or research ethics committees at the University of California San Francisco (USA), University of Heidelberg (Germany), De La Salle Medical and Health Sciences Institute (Philippines), Uganda National Council for Science and Technology (Uganda), Hanoi Lung Hospital (Vietnam), and University of Zambia (Zambia).

## Results

Across the four high-burden countries, tongue swab-based molecular testing using MiniDock MTB was associated with substantially lower per-test costs compared to sputum-based testing with Xpert Ultra (Table 1). Absolute cost reductions ranged from USD 12.55 to USD 17.85, representing a 47.8–55.1% decrease. These differences were primarily driven by lower device and cartridge costs for MiniDock MTB relative to Xpert Ultra. Per-test costs for MiniDock MTB using sputum swabs were only marginally higher than for tongue swabs (Table 1).

**Table 1:** Unit cost estimates for sputum Xpert Ultra. tongue swab and sputum swab MiniDock MTB testing (2024 USD)

| Cost item | Philippines | Uganda | Vietnam | Zambia |
| --- | --- | --- | --- | --- |
| **Sputum Xpert Ultra** |  |  |  |  |
| Clinic staff time: sample collection | 3.40 (1.98–5.37) | 4.14 (2.90–5.40) | 2.73 (2.26–3.28) | 1.69 (0.86–2.80) |
| Consumables for sample collection (sputum cup) | 0.16 (0.09–0.22) | 0.11 (0.10–0.12) | 0.28 (0.09–0.45) | 0.64 (0.14–1.16) |
| PPE per encounter (mask, gloves, disinfectant) | 1.09 (0.96–1.37) | 0.28 (0.20–0.45) | 0.41 (0.27–0.74) | 0.89 (0.31–1.49) |
| Laboratory cost (staff time, consumables, recordkeeping) | 17.77 (14.56–20.97) | 7.52 (5.12–9.86) | 10.00 (–) | 20.34 (19.05–21.50) |
| Xpert Ultra cartridge | * | 9.00 (8.06–9.95) | 12.98 (11.08–14.91) | 8.78 (6.62–10.97) |
| GeneXpert instrument amortization (per test) | 0.46 (0.42–0.50) | 0.46 (0.42–0.50) | † | † |
| Annual calibration and maintenance | 0.03 (–) | 0.03 (–) | † | † |
| **Total cost per person tested** | **22.88 (18.97–26.67)** | **21.50 (18.52–24.59)** | **26.40 (24.33–28.52)** | **32.34 (29.37–35.39)** |
| **Tongue swab MiniDock MTB** |  |  |  |  |
| Clinic staff time: sample collection | 2.98 (1.83–4.48) | 4.01 (2.80–5.16) | 1.69 (1.09–2.32) | 1.60 (0.75–2.70) |
| Consumables for sampling and processing (Pluslife Test Kit) | 4.00 (–) | 4.00 (–) | 4.00 (–) | 4.00 (–) |
| PPE per encounter (mask, gloves, disinfectant) | 1.09 (0.96–1.37) | 0.28 (0.20–0.45) | 0.41 (0.27–0.74) | 0.89 (0.31–1.49) |
| Laboratory cost (staff time, consumables, recordkeeping) | 3.94 (3.36–4.49) | 5.08 (4.60–5.54) | 4.51 (3.08–5.91) | 8.00 (–) |
| MiniDock MTB instrument amortization (per test) | 0.35 (–) | 0.35 (–) | 0.35 (–) | † |
| **Total cost per person tested** | **12.36 (11.01–14.01)** | **13.73 (12.39–14.95)** | **10.96 (9.29–12.65)** | **14.49 (13.32–15.85)** |
| **Sputum Swab MiniDock MTB** |  |  |  |  |
| Clinic Staff Time: Sample collection | 3.40 (1.98;5.37) | 4.14 (2.90;5.40) | 2.73 (2.26;3.28) | 1.69 (0.86;2.80) |
| Consumables for Sample Collection | 0.16 (0.09;0.22) | 0.11 (0.10;0.12) | 0.28 (0.09;0.45) | 0.64 (0.14;1.16) |
| PPE per encounter (mask, gloves, disinfectant) | 1.09 (0.96;1.37) | 0.28 (0.20;0.45) | 0.41 (0.27;0.74) | 0.89 (0.31;1.49) |
| Laboratory cost (staff time, consumables for testing and recording) | 4.00 (3.40;4.57) | 5.20 (4.70;5.67) | 4.51 (3.08;5.91) | 8.00 (-) |
| MiniDock MTB sampling consumables and test chip | 4.00 (-) | 4.00 (-) | 4.00 (-) | 4.00 (-) |
| MiniDock MTB instrument amortization (per test) | 0.35 (-) | 0.35 (-) | † | † |
| **Total Costs per person tested** | **12.99 (11.40;14.99)** | **14.07 (12.67;15.54)** | **11.92 (10.33;13.67)** | **15.22 (13.88;16.70)** |

**Notes:**
Costs are presented per patient tested and include staff time, sample and test consumables, laboratory processing, and device acquisition and maintenance. All costs are reported in 2024 USD. Overhead costs were excluded, as relative differences between sampling methods were negligible. Uncertainty ranges (in parentheses) represent 95% uncertainty intervals. Clinic staff time includes time spent by clinical personnel on sample collection (nurse) and consultation (clinician/doctor), calculated as salary per minute multiplied by activity duration. Time estimates were obtained from a healthcare worker survey conducted within the TSwaY study. A 5% adjustment was applied to account for training.
The Pluslife Test Kit includes the swab, buffer, collection tube, test chip (MiniDock MTB Test cartridge), and hazard bag. Information was obtained directly from the manufacturer.
PPE costs per encounter include single-use items (e.g., gloves, disinfectant) and shared-use items (e.g., gowns, masks), costed per shift and proportionally allocated across patients tested per day. Cost data were obtained from supplier quotes, donation records, and procurement departments.
Laboratory costs include technician time, consumables (e.g., gloves, masks, disinfectant), and laboratory overheads.
Device amortization and maintenance reflect acquisition, warranty, and maintenance costs of the testing platform, apportioned per test over the expected lifespan. Estimates were derived from the Stop TB Partnership product catalogues or directly from manufacturers. The Pluslife amortization estimate already includes maintenance.
\*Xpert Ultra Cartridge in the Philippines is included in the laboratory cost.
† Device amortization and maintenance costs for sputum testing in Zambia and Vietnam were included within laboratory costs and are not shown separately.

Under the current standard of care, sputum Xpert Ultra yielded 67 (95% UI: 53 to 80) bacteriologically confirmed TB diagnoses at an estimated cost of USD 528 (95%UI: 424 to 687) per diagnosis. This strategy was extendedly dominated by alternatives incorporating tongue swab MiniDock MTB.

Full replacement of sputum Xpert Ultra with tongue swab MiniDock MTB (*tongue swab-only*) was the least costly approach but yielded the fewest bacteriologically confirmed TB diagnoses: 62 (95% UI: 48 to 75) at a cost of USD 300 (95% UI: 244 to 388) per diagnosis (Table 2). The *limited combined* strategy yielded 12 (95% UI: -1 to 22) additional diagnoses at an incremental cost of USD 1,507 per additional diagnosis compared with *tongue swab-only*. When compared directly with sputum Xpert Ultra (standard of care), the limited combined strategy yielded 7 (95% UI: 2 to 12) additional diagnoses at an incremental cost of USD 198 (95% UI 108 to 469) per diagnosis (*Supplementary Table S2*). The *extended combined* strategy yielded 15 (95% UI: 8 to 22) additional diagnoses at an incremental cost of USD 1,004 (95% UI: 661 to 1,655) per additional diagnosis compared with the *limited combined* strategy (Table 2).

**Table 2.** Cost, number of TB diagnoses, and incremental cost-effectiveness ratios (ICERs) for three scenarios integrating tongue swab-based testing using MiniDock MTB into primary healthcare, alongside sputum Xpert Ultra (standard of care). Point estimates for TB diagnoses are based on observed data from the TSwaY study. Values in brackets indicate 95% uncertainty intervals, derived from probabilistic sensitivity analysis. Participant totals are listed beneath each country name. All incremental estimates are calculated relative to the next least costly, non-dominated strategy.

| Setting/strategy | Total cost<br>(Thousand USD) | TB diagnoses | Incremental cost<br>(Thousand USD) | Incremental TB<br>diagnoses | ICER (USD per<br>additional<br>TB diagnosis) |
| --- | --- | --- | --- | --- | --- |
| **All countries**<br>**(n = 1370)** |  |  |  |  |  |
| TS-only (MiniDock MTB) | 18.7 (17.3; 19.9) | 62 (48; 75) | - | - | (Reference) |
| Sputum Xpert Ultra (SoC) | 35.4 (33.4; 37.1) | 67 (53; 80) | 16.7 (14.5; 18.5) | 5 (-10; 16) | (Ext. dominated)* |
| Limited combined | 36.7 (34.8; 38.5) | 74 (58; 87) | 18.1 (15.9; 19.9) | 12 (-1; 22) | 1507† |
| Extended combined | 51.8 (49.2; 54.0) | 89 (72; 104) | 15.1 (13.9; 16.0) | 15 (8; 22) | 1004 (661; 1655) |
| **Philippines**<br>**(n = 182)** |  |  |  |  |  |
| TS-only (MiniDock MTB) | 2.5 (1.8; 2.5) | 12 (6; 18) | - | - | (Reference) |
| Sputum Xpert Ultra (SoC) | 4.1 (3.4; 4.6) | 14 (8; 20) | 1.6 (1.2; 2.5) | 2 (-6; 9) | (Ext. dominated)* |
| Limited combined | 4.4 (3.7; 4.9) | 19 (11; 26) | 1.9 (1.5; 2.8) | 7 (1; 13) | 276 (133; 927) |
| Extended combined | 6.2 (5.2; 6.5) | 21 (13; 29) | 1.8 (1.2; 2.5) | 2 (0; 5) | 903 (282; 1647) |
| **Vietnam**<br>**(n = 626)** |  |  |  |  |  |
| TS-only (MiniDock MTB) | 7.3 (6.2; 8.3) | 16 (9; 23) | - | - | (Reference) |
| Sputum Xpert Ultra (SoC) | 16.3 (15.0; 17.4) | 21 (12; 29) | 9.0 (7.3; 10.6) | 5 (-2; 11) | 1805† |
| Limited combined | 16.4 (15.1; 17.5) | 21 (12; 29) | 0.1 (0.1; 0.1) | 0 (0; 0) | (Dominated) |
| Extended combined | 22.9 (21.2; 24.5) | 25 (16; 34) | 6.7 (5.6; 7.4) | 4 (1; 7) | 1665 (810; 6247) |
| **Uganda**<br>**(n = 218)** |  |  |  |  |  |
| TS-only (MiniDock MTB) | 3.1 (2.8; 3.4) | 3 (0; 6) | - | - | (Reference) |
| Sputum Xpert Ultra (SoC) | 4.9 (4.3; 5.4) | 6 (2; 10) | 1.8 (1.3; 2.3) | 3 (0; 6) | 606 (238; 1954) |
| Limited combined | 5.4 (4.8; 6.0) | 6 (2; 10) | 2.4 (1.8; 2.8) | 0 (0; 0) | (Dominated) |
| Extended combined | 7.9 (7.1; 8.5) | 6 (2; 10) | 2.4 (2.2; 2.6) | 0 (0; 0) | (Dominated) |
| **Zambia**<br>**(n = 344)** |  |  |  |  |  |
| TS-only (MiniDock MTB) | 5.9 (5.3; 6.6) | 31 (21; 40) | - | - | (Reference) |
| Sputum Xpert Ultra (SoC) | 10.1 (9.2; 11.0) | 26 (17; 34) | 4.2 (3.3; 5.0) | -5 (-13; 2) | (Dominated) |
| Limited combined | 10.6 (9.7; 11.5) | 28 (19; 37) | 4.7 (3.8; 5.5) | -3 (-11; 3) | (Dominated) |
| Extended combined | 14.8 (13.5; 15.9) | 37 (27; 46) | 8.9 (7.8; 9.7) | 6 (2; 10) | 1475 (797; 4444) |

TS = tongue swab, SoC = standard of care, USD = US dollars, ICER = incremental cost-effectiveness ratio, Ext. dominated = extendedly dominated.
\* Sputum Xpert Ultra was extendedly dominated, meaning it was more costly and yielded fewer bacteriologically confirmed TB diagnoses than at least one combination of the other evaluated strategies. Strategies incorporating tongue swab-based testing using MiniDock MTB – either alone or in combination with sputum Xpert Ultra – achieved higher diagnostic yield at lower or comparable cost.
† The uncertainty interval for this ICER could not be calculated because incremental diagnoses were negative in some model iterations.

At lower WTP thresholds, the *tongue swab-only* strategy had the highest probability of yielding the greatest NMB (Figure 1a). As WTP increased, this probability declined, and the *extended combined* strategy became the most cost-effective option beyond USD 1,256 per diagnosis. When the *tongue swab-only* strategy was excluded from this analysis – given its lower diagnostic yield than the standard of care – the *limited combined* strategy had the highest probability of being optimal between USD 200 and USD 1,000 per diagnosis (Figure 1b). Across all WTP thresholds, sputum Xpert Ultra alone was consistently outperformed by at least one tongue swab-inclusive strategy (Figure 1a,b).

**Figure 1.**
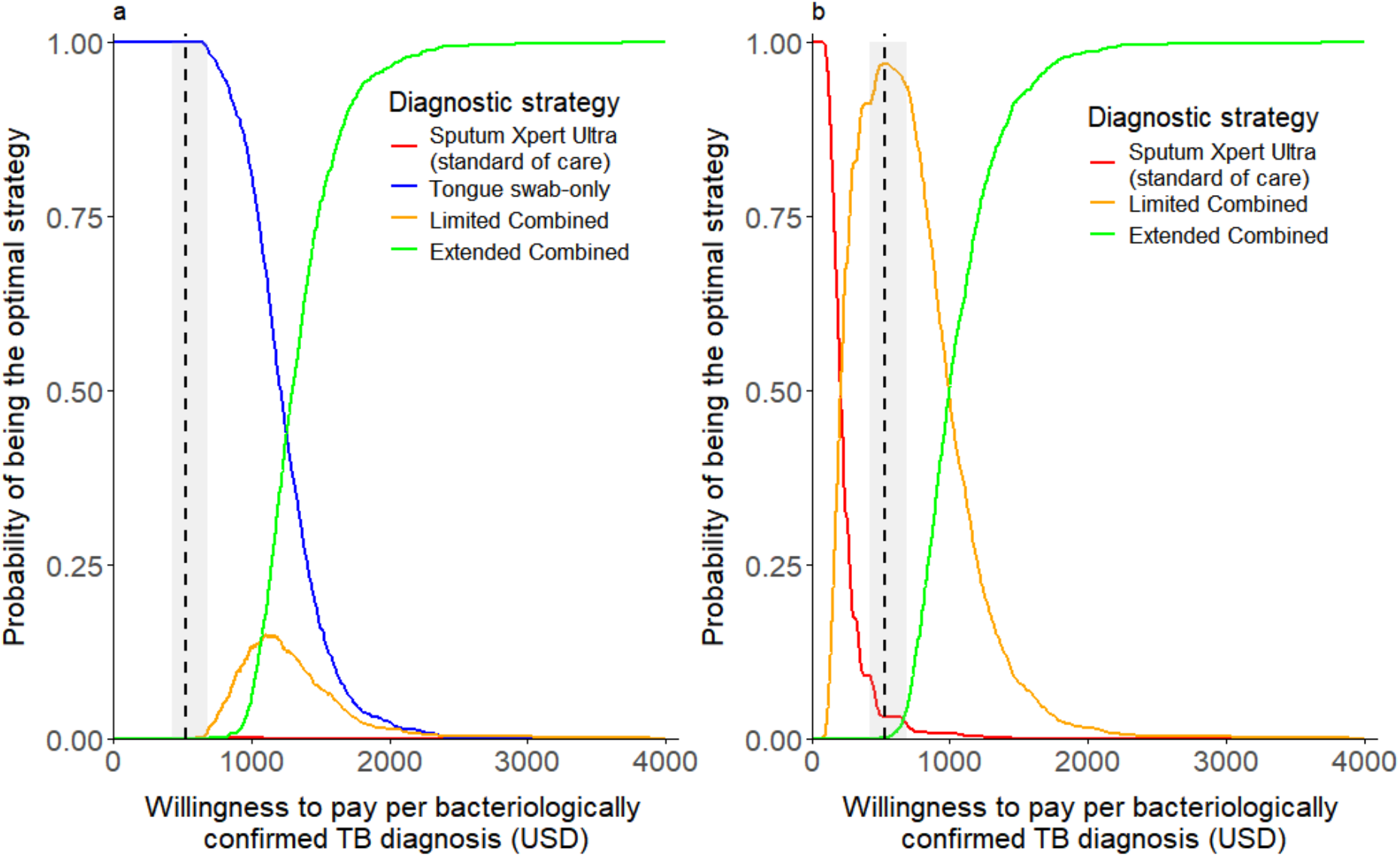
Cost-effectiveness acceptability curves for four tuberculosis (TB) diagnostic strategies. Curves show the probability of each strategy being optimal across a range of willingness-to-pay (WTP) thresholds per additional bacteriologically confirmed TB diagnosis. The optimal strategy at each WTP is defined as the one with the highest expected net monetary benefit, based on probabilistic analysis. **Panel 1a:** All four strategies included: (1) *sputum Xpert Ultra* (standard of care), (2) *tongue swab-only* (MiniDock MTB), (3) *limited combined* strategy (sputum Xpert Ultra plus tongue swab MiniDock MTB for sputum-scarce individuals), and (4) *extended combined* strategy (sputum Xpert Ultra plus MiniDock MTB for sputum-scarce individuals and those with negative or indeterminate sputum results). **Panel 1b:** Comparison excluding the tongue swab-only strategy, which yielded fewer diagnoses than the standard of care. The vertical dashed line indicates the estimated cost per diagnosis under the standard of care (USD 528), used as a benchmark WTP threshold.

Country-level results varied due to differences in per-test costs and diagnostic yield (Table 2; *Supplementary Figures S1 and S2*).

Incorporating the simulated *integrated combined* strategy, in which MiniDock MTB was used to test either a sputum swab (if available) or a tongue swab for sputum-scarce individuals, yielded 8 (95% UI: –4 to 19) additional diagnoses at an incremental cost of USD 165 per additional diagnosis compared with the *tongue swab-only* strategy. This strategy strictly dominated the *standard of care* (sputum Xpert Ultra) which was more costly and yielded fewer diagnosis than the *integrated combined* strategy (Table 3). Across all WTP thresholds, single-platform MiniDock MTB strategies consistently outperformed those involving Xpert Ultra. The tongue swab-only strategy had the highest probability of yielding the greatest net monetary benefit at WTP values up to USD 161, while the integrated combined strategy became optimal at higher thresholds (Figure 2a). If the diagnostic yield of sputum swab testing were assumed to match that of sputum Xpert Ultra, the threshold at which the integrated combined strategy became optimal would fall to USD 115 (Figure 2b).

**Table 3.** Cost, number of TB diagnoses, and incremental cost-effectiveness ratios (ICERs) for the extended analysis including the simulated *integrated combined* strategy. The *integrated combined* strategy assumes MiniDock MTB testing of sputum swabs for individuals able to produce sputum and tongue swabs for those unable to do so. Point estimates for TB diagnoses are based on observed data from the TSwaY study. Values in brackets indicate 95% uncertainty intervals derived from probabilistic sensitivity analysis. Participant totals are listed beneath each country name. All incremental estimates are calculated relative to the next least costly, non-dominated strategy.

| Setting/strategy | Total cost<br>(Thousand USD) | TB diagnoses | Incremental cost<br>(Thousand USD) | Incremental TB<br>diagnoses | ICER (USD per<br>additional<br>TB diagnosis) |
| --- | --- | --- | --- | --- | --- |
| **All countries**<br>**(n = 1370)** |  |  |  |  |  |
| TS-only (MiniDock MTB) | 18.7 (17.3; 19.9) | 62 (48; 75) | - | - | (Reference) |
| Integrated combined | 20.0 (18.6; 21.3) | 70 (54; 86) | 1.3 (0.7; 1.8) | 8 (-4; 19) | 165<sup>†</sup> |
| Sputum Xpert Ultra (SoC) | 35.4 (33.4; 37.1) | 67 (53; 80) | 15.5 (13.3; 17.2) | -3 (-9; 1) | (Dominated) |
| Limited combined | 36.7 (34.8; 38.5) | 74 (58; 87) | 16.7 (14.8; 18.6) | 4 (-20; 22) | 4191<sup>†</sup> |

TS = tongue swab, SoC = standard of care, USD = US dollars, ICER = incremental cost-effectiveness ratio, Ext. dominated = extendedly dominated.
<sup>†</sup> The uncertainty interval for this ICERs could not be calculated because incremental diagnoses were negative in some model iterations.

**Figure 2.**
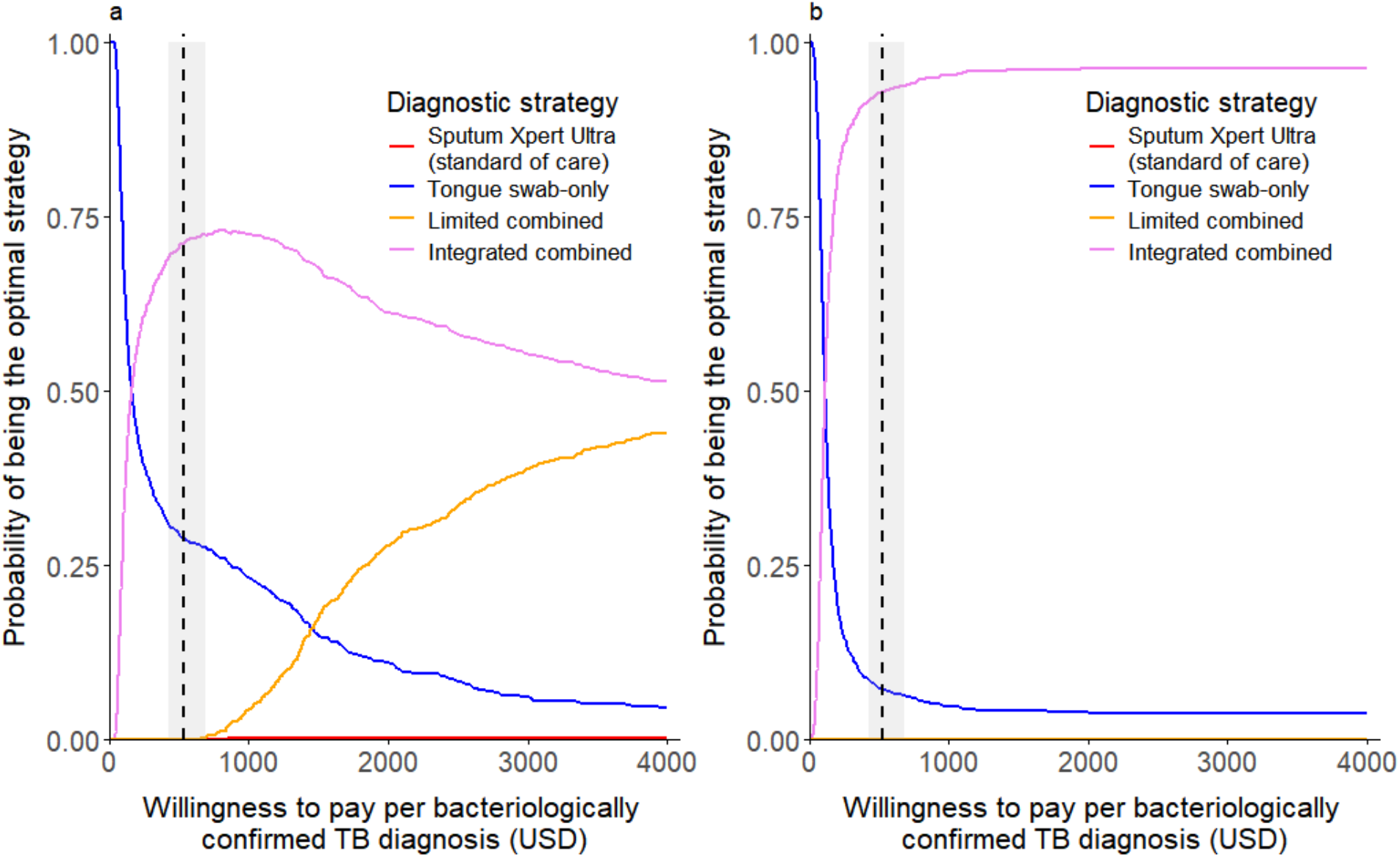
Cost-effectiveness acceptability curves including the simulated integrated combined strategy using MiniDock MTB. The *integrated combined* strategy assumes MiniDock MTB testing of sputum swabs for individuals able to produce sputum and tongue swabs for those unable to do so. Curves show the probability of each strategy being cost-effective across a range of willingness-to-pay (WTP) thresholds per additional bacteriologically confirmed TB diagnosis. The optimal strategy at a given WTP is defined as the one with the highest expected net monetary benefit (NMB), based on probabilistic sensitivity analysis. **Panel 2a:** Comparison of all evaluated strategies except for the *extended combined* strategy. **Panel 2b:** Same comparison as in 2a, but assuming the diagnostic yield of sputum swab MiniDock MTB matched that of sputum Xpert Ultra (i.e. relative yield = 1). The vertical dashed line indicates the estimated cost per diagnosis under the standard of care (USD 528), used as a benchmark WTP threshold.

Sensitivity analysis in which we assumed that all sputum samples were tested did not meaningfully change the results (*Supplementary Table S3*).

## Discussion

In this study, we estimated the cost and cost-effectiveness of integrating tongue swab MiniDock MTB testing into routine TB diagnostic services in four high-burden countries. Our analysis found that MiniDock MTB testing using tongue swabs is more affordable than sputum Xpert Ultra, driven by lower costs for devices, cartridges, and maintenance. When used in combination with sputum Xpert Ultra, tongue swab testing increased diagnostic yield at reasonable additional cost. Across all WTP thresholds per bacteriologically confirmed TB diagnosis, sputum Xpert Ultra – the current standard of care – was consistently outperformed by strategies incorporating tongue swab MiniDock MTB.

We found that replacing sputum Xpert Ultra entirely with tongue swab MiniDock MTB (*tongue swab-only*) would be cost-saving but resulted in slightly fewer TB diagnoses. Combined testing strategies, where sputum Xpert Ultra remains the primary test and tongue swab testing is added for those unable to produce sputum (*limited combined*), or additionally for those with negative or indeterminate sputum results (*extended combin*ed), increased the number of bacteriologically confirmed TB diagnoses at incremental costs that fall within widely accepted cost-effectiveness thresholds. The extended combined strategy had the highest probability of being optimal at WTP values above USD 1,256 per additional diagnosis. Although this exceeds the current cost per diagnosis, it remains below or within the range of per-capita GDP in all four countries (USD 1,073–4,717) and well within established thresholds for cost-effectiveness.^22^ Assuming each bacteriologically confirmed TB diagnosis averts 2–3 disability-adjusted life years (DALYs)^23^, the corresponding WTP threshold would fall well below 0.5x GDP per capita in all settings.

To examine the potential of a streamlined, single-platform diagnostic workflow, we incorporated a simulated “*integrated combined*” strategy that would use MiniDock MTB to test sputum swabs as the primary sample and tongue swabs for those unable to produce sputum. This strategy increased the number of confirmed diagnoses at minimal additional cost compared to tongue swab-only testing and strictly dominated the standard of care. Notably, MiniDock MTB-based one-platform strategies – either *tongue swab-only* or *integrated combined* – outperformed all strategies that included Xpert Ultra at any WTP threshold. These findings support the potential cost-effectiveness and operational feasibility of consolidating TB molecular testing on a single, portable platform, offering particular advantages for the decentralization of care.

A key strength of this analysis is its use of empirical diagnostic and costing data, allowing for context-specific estimates with high relevance to national TB programmes. However, several limitations must be acknowledged. First, the TSwaY study did not include testing against a microbiological gold standard such as culture. As such, some diagnoses may have been false positives. However, this is unlikely to substantially affect our conclusions, given the high specificity of both Xpert Ultra and MiniDock MTB in prior evaluations. Second, all sampling in TSwaY was performed by trained research staff, and the yield of tongue swabs in routine care could vary. On the other hand, given known challenges with sputum collection in real-world settings, tongue swab testing may perform even better operationally than estimated here. Third, we assumed onsite testing for both modalities, which does not fully account for potential operational advantages of tongue swab testing – especially its suitability for decentralized use. If confirmed, these advantages could further reduce loss to follow-up and delays in treatment initiation. Fourth, our analysis was limited to bacteriologically confirmed TB and did not consider broader clinical outcomes, including empiric treatment. Lastly, we adopted a healthcare system perspective and did not account for patient-incurred costs.

In conclusion, our health-economic evaluation provides robust support for integrating tongue swab and sputum swab MiniDock MTB testing into TB diagnostic pathways in high-burden settings. Tongue swab-based molecular testing can serve as a practical and cost-effective complement to sputum-based testing, with added operational flexibility. An integrated strategy using MiniDock MTB for both specimen types could further streamline workflows, reduce infrastructure needs, and improve access to timely diagnosis. Future research should evaluate the performance and feasibility of such approaches in diverse populations and settings, including children, people living with HIV, and lower-burden areas. With growing momentum toward decentralized and person-centered TB care, tongue swab-based diagnostics – alone or integrated with sputum-based diagnostics – may play an important role in expanding access to effective TB testing globally.

## Data Availability

All data required to reproduce this health-economic analysis are provided in this manuscript and the supplementary material. De-identified participant data from the TSwaY principal study that underlie the results reported in this article, study protocol, statistical analytical plan, and/or analytic code are available upon reasonable request from researchers with a methodologically sound proposal. Proposals should be directed to; to gain access, data requestors will need to sign a data access agreement.

## Acknowledgments

We gratefully acknowledge the individuals who participated in the TSwaY study and also extend our sincere thanks to the administrative teams and clinical staff at the participating health centers for their support and dedication. We thank the TSwaY study team for supporting this analysis. We are grateful to David Dowdy and Jason Andrews for helpful feedback on the manuscript.

## Data availability statement

All data required to reproduce this health-economic analysis are provided in this manuscript and the supplementary material. De-identified participant data from the TSwaY study that underlie the results reported in this article, study protocol, statistical analytical plan, and/or analytic code to researchers who provide a methodologically sound proposal. Proposals should be directed to; to gain access, data requestors will need to sign a data access agreement.

## References

1. Albert H, Nathavitharana RR, Isaacs C, Pai M, Denkinger CM, Boehme CC. Development, roll-out and impact of Xpert MTB/RIF for tuberculosis: wh at lessons have we learnt and how can we do better? European Respiratory Journal 2016; 48(2): 516–25.

2. Creswell J, Codlin AJ, Andre E, et al. Results from early programmatic implementation of Xpert MTB/RIF testin g in nine countries. BMC Infectious Diseases 2014: 2.

3. Naidoo P, Dunbar R, Lombard C, et al. Comparing Tuberculosis Diagnostic Yield in Smear/Culture and Xpert® MT B/RIF-Based Algorithms Using a Non-Randomised Stepped-Wedge Design. PloS one 2016; 11(3): e0150487.

4. Boum Y, 2nd, Orikiriza P, Rojas-Ponce G, et al. Use of colorimetric culture methods for detection of Mycobacterium tuberculosis complex isolates from sputum samples in resource-limited settings. J Clin Microbiol 2013; 51(7): 2273–9.

5. Divala TH, Corbett EL, Kandulu C, et al. Trial-of-antibiotics to assist tuberculosis diagnosis in symptomatic adults in Malawi (ACT-TB study): a randomised controlled trial. Lancet Glob Health 2023; 11(4): e556–e65.

6. Grant AD, Charalambous S, Tlali M, et al. Algorithm-guided empirical tuberculosis treatment for people with advanced HIV (TB Fast Track): an open-label, cluster-randomised trial. Lancet HIV 2020; 7(1): e27–e37.

7. Broger T, Koeppel L, Huerga H, et al. Diagnostic yield of urine lipoarabinomannan and sputum tuberculosis tests in people living with HIV: a systematic review and meta-analysis of individual participant data. The Lancet Global Health 2023; 11(6): e903–e16.

8. Broger T, Marx FM, Theron G, et al. Diagnostic yield as an important metric for the evaluation of novel tuberculosis tests: rationale and guidance for future research. The Lancet Global Health 2024; 12(7): e1184–e91.

9. Nathavitharana RR, Garcia-Basteiro AL, Ruhwald M, Cobelens F, Theron G. Reimagining the status quo: How close are we to rapid sputum-free tube rculosis diagnostics for all? eBioMedicine 2022; 78: 103939.

10. Manoj Kumar K, Borkman A, Kim A, et al. Preferences for Tongue Swab versus Sputum Collection for Tuberculosis Testing: A Multi-Country Survey. medRxiv 2025: 2025.07.04.25330895.

11. Sales A, Le H, Muzazu S, et al. “That is why I trust”: A qualitative study on acceptability and feasibility of novel tongue swab diagnostics to assess people presenting with tuberculosis symptoms in Viet Nam and Zambia. medRxiv 2025: 2025.06.30.25330535.

12. Wood RC, Luabeya AK, Dragovich RB, et al. Diagnostic accuracy of tongue swab testing on two automated tuberculos is diagnostic platforms, Cepheid Xpert MTB/RIF Ultra and Molbio Truena t MTB Ultima. Journal of Clinical Microbiology 2024; 62(4).

13. Church EC, Steingart KR, Cangelosi GA, Ruhwald M, Kohli M, Shapiro AE. Oral swabs with a rapid molecular diagnostic test for pulmonary tuberculosis in adults and children: a systematic review. The Lancet Global Health 2024; 12(1): e45–e54.

14. Steadman A, Kumar KM, Asege L, et al. Diagnostic accuracy of swab-based molecular tests for tuberculosis usi ng novel near point-of-care platforms: A multi-country evaluation. 2025.

15. Mycobacterium Tuberculosis(Sputum/Tongue swab sample)-Pluslife. Accessed August 03, 2025. https://www.pluslife.com/productinfo/1268173.html.

16. Schünemann HJ, Mustafa R, Brozek J, et al. GRADE Guidelines: 16. GRADE evidence to decision frameworks for tests in clinical practice and public health. J Clin Epidemiol 2016; 76: 89–98.

17. Moe CA, Luswata R, Barrameda J, et al. Diagnostic yield of tongue swab-compared to sputum-based molecular te sting for tuberculosis in four high-burden countries. 2025.

18. Husereau D, Drummond M, Augustovski F, et al. Consolidated Health Economic Evaluation Reporting Standards 2022 (CHEERS 2022) Statement: Updated Reporting Guidance for Health Economic Evaluations. Value in health : the journal of the International Society for Pharmacoeconomics and Outcomes Research 2022; 25(1): 3–9.

19. Steadman A, Kumar KM, Asege L, et al. Diagnostic accuracy of swab-based molecular tests for tuberculosis using novel near point-of-care platforms: A multi-country evaluation. medRxiv 2025: 2025.04.12.25325603.

20. International Monetary Fund (IMF). Exchange rates [Internet]. Washington (DC): IMF; 2024. Available from: https://data.imf.org.

21. Briggs AH, Wonderling DE, Mooney CZ. Pulling cost-effectiveness analysis up by its bootstraps: a non-parametric approach to confidence interval estimation. Health Econ 1997; 6(4): 327–40.

22. Woods B, Revill P, Sculpher M, Claxton K. Country-Level Cost-Effectiveness Thresholds: Initial Estimates and the Need for Further Research. Value in Health 2016; 19(8): 929–35.

23. Vassall A, van Kampen S, Sohn H, et al. Rapid diagnosis of tuberculosis with the Xpert MTB/RIF assay in high burden countries: a cost-effectiveness analysis. PLoS Med 2011; 8: e1001120.

