## Supplementary Information for "Cost and cost-effectiveness of swab-based molecular testing for tuberculosis in the Philippines, Uganda, Vietnam, and Zambia"

**Simulation of diagnostic yield under the integrated combined strategy (sputum swab and tongue swab with MiniDock MTB)**

We simulated the diagnostic yield of an integrated combined strategy in which both, sputum swabs and tongue swabs, are tested using MiniDock MTB. Operationally, sputum would be requested from all participants. For individuals able to produce sputum, a swab would be dipped into the sputum specimen and tested on MiniDock MTB. Those unable to produce sputum would instead undergo tongue swab sampling for MiniDock MTB testing.

The diagnostic yield of sputum swab MiniDock MTB was estimated using a relative yield approach, defined as the probability of a positive result on sputum swab MiniDock MTB among individuals with a positive result on sputum Xpert Ultra. This relative yield, denoted as *R*, was estimated from paired data from two independent evaluations of the MiniDock MTB platform using sputum swabs (sources: see main manuscript). In these evaluations, 62 of 64 and 285 of 304 individuals who tested positive on sputum Xpert Ultra also tested positive on MiniDock MTB using a sputum swab. These data were pooled, resulting in a total of 347 tongue swab MiniDock MTB-positive cases out of 368 sputum Xpert Ultra-positive individuals.

To propagate uncertainty in this estimate, we placed a non-informative Jeffreys prior (*Beta[0.5, 0.5]*) on the binomial distribution and derived the posterior as:

$$R \mid data \sim Beta(347 + 0.5,\ 21 + 0.5)$$

The number of individuals testing positive with sputum swab MiniDock MTB in iteration m was calculated in the Monte Carlo simulation by multiplying, at each iteration, a sampled relative yield estimate *R* with the previously sampled number of individuals testing positive on sputum Xpert Ultra.

This approach allowed us to incorporate uncertainty in both the yield of sputum Xpert Ultra (from TSwaY) and the relative yield of sputum swab MiniDock MTB (from external data). We report the point estimate as the mean across 1,000 iterations, with 95% uncertainty intervals defined by the 2.5th and 97.5th percentiles of the resulting distribution.

**Table S1. Cross-tabulation of sputum Xpert Ultra and tongue swab MiniDock MTB Test results from the TSwaY study.**

The table presents the distribution of diagnostic outcomes by sample type, showing the number of individuals with each combination of sputum and tongue swab results. Result categories are defined as follows: sample not provided: no sample collected; no result: sample not processed or yielded an indeterminate result; negative: includes negative results and trace (Xpert Ultra only); positive: bacteriologically confirmed positive result.

| | | **TONGUE SWAB MiniDock MTB** | | | | |
|---|---|---|---|---|---|---|
| | | Sample not provided | No result | TS negative | TS positive | **Total** |
| **SPUTUM Xpert Ultra** | Sample not provided | 0 | 0 | 104 | 7 | **111** |
| | No result | 0 | 0 | 49 | 3 | **52** |
| | Negative | 1 | 24 | 1103 | 12 | **1140** |
| | Positive | 1 | 1 | 25 | 40 | **67** |
| | **Total** | **2** | **25** | **1281** | **62** | **1370** |

**Table S2. Cost, number of TB diagnoses, and incremental cost-effectiveness ratios (ICERs) for two combined strategies integrating tongue swab-based testing using MiniDock MTB into primary healthcare alongside sputum Xpert Ultra (standard of care), excluding the tongue swab-only strategy.**

Point estimates for TB diagnoses are based on observed data from the TSwaY study. Values in brackets indicate 95% uncertainty intervals, derived from probabilistic sensitivity analysis. Participant totals are listed beneath each country name. All incremental estimates are calculated relative to the next least costly, non-dominated strategy.

| Setting/strategy | Total cost (Thousand USD) | TB diagnoses | Incremental cost (Thousand USD) | Incremental TB diagnoses | ICER (USD per additional TB diagnosis)* |
|---|---|---|---|---|---|
| **All countries (n = 1370)** | | | | | |
| Sputum Xpert Ultra (SoC) | 35.4 (33.4; 37.1) | 67 (53; 80) | - | - | Reference |
| Limited combined | 36.7 (34.8; 38.5) | 74 (58; 87) | 1.4 (1.3; 1.5) | 7 (2; 12) | 198 (108; 469) |
| Extended combined | 51.8 (49.2; 54.0) | 89 (72; 104) | 15.1 (13.9; 16.0) | 15 (8; 22) | 1004 (661; 1655) |

**Table S3. Sensitivity analysis assuming successful processing of all tongue swab and sputum samples (i.e. no missing samples not processed): Cost, number of TB diagnoses, and incremental cost-effectiveness ratios (ICERs) for onsite sputum Xpert Ultra testing (standard of care) and three MiniDock MTB tongue swab-based testing strategies.** Point estimates for TB diagnoses are based on observed data from the TSwaY study. Values in brackets indicate 95% uncertainty intervals, derived from probabilistic sensitivity analysis. Participant totals are listed beneath each country name. All incremental estimates are calculated relative to the next least costly, non-dominated strategy.

| Setting/strategy | Total cost (Thousand USD) | TB diagnoses | Incremental cost (Thousand USD) | Incremental TB diagnoses | ICER (USD per additional TB diagnosis) |
|---|---|---|---|---|---|
| **All countries (n = 1370)** | | | | | |
| TS-only (MiniDock MTB) | 18.7 (17.3; 19.9) | 63 (49; 76) | - | - | (Reference) |
| Sputum Xpert Ultra (SoC) | 35.4 (33.4; 37.1) | 70 (55; 83) | 16.7 (14.4; 18.5) | 6.7 (-9; 18) | (Ext. dominated*) |
| Limited combined | 36.7 (34.7; 38.4) | 77 (63; 92) | 18.1 (15.8; 19.9) | 13.7 (1; 28) | 1318 (554; 3116) |
| Extended combined | 51.8 (49.2; 54.0) | 92 (75; 109) | 15.0 (13.9; 16.0) | 15.4 (3; 21) | 976 (674; 2855) |
| **Philippines (n = 182)** | | | | | |
| TS-only (MiniDock MTB) | 2.5 (1.8; 2.5) | 12 (6; 18) | - | - | (Reference) |
| Sputum Xpert Ultra (SoC) | 4.1 (3.4; 4.6) | 14 (8; 20) | 1.6 (1.0; 2.1) | 2 (-6; 9) | (Ext. dominated*) |
| Limited combined | 4.4 (3.7; 4.9) | 19 (13; 25) | 1.9 (1.4; 2.5) | 7 (-1; 14) | 276† |
| Extended combined | 6.2 (5.2; 6.5) | 21 (13; 29) | 1.8 (1.0; 2.1) | 2 (-2; 7) | 903† |
| **Vietnam (n = 626)** | | | | | |
| TS-only (MiniDock MTB) | 7.3 (6.2; 8.3) | 16 (9; 24) | - | - | (Reference) |
| Sputum Xpert Ultra (SoC) | 16.3 (15.0; 17.4) | 21 (12; 29) | 9.0 (7.3; 10.6) | 5 (-2; 11) | 1,918† |
| Limited combined | 16.4 (15.1; 17.5) | 21 (12; 29) | 9.1 (7.4; 10.7) | 0 (-) | (Dominated) |
| Extended combined | 22.9 (21.2; 24.5) | 25 (16; 34) | 6.5 (21.1; 24.5) | 4 (1; 7) | 1595 (721; 6189) |
| **Uganda (n = 218)** | | | | | |
| TS-only (MiniDock MTB) | 3.1 (2.8; 3.4) | 3 (0; 6) | - | - | (Reference) |
| Sputum Xpert Ultra (SoC) | 4.9 (4.3; 5.4) | 6 (2; 10) | 1.8 (1.3; 2.3) | 3 (0; 6) | 606 (238; 1954) |
| Limited combined | 5.4 (4.8; 6.0) | 6 (2; 10) | 2.4 (1.8; 2.8) | 0 (-) | (Dominated) |
| Extended combined | 7.9 (7.1; 8.5) | 6 (2; 10) | 2.4 (2.2; 2.6) | 0 (-) | (Dominated) |
| **Zambia (n = 344)** | | | | | |
| TS-only (MiniDock MTB) | 5.9 (5.3; 6.6) | 32 (21; 41) | - | - | (Reference) |
| Sputum Xpert Ultra (SoC) | 10.1 (9.2; 11.0) | 31 (19; 40) | 4.2 (3.3; 5.0) | -1 (-10; 7) | (Dominated) |
| Limited combined | 10.6 (9.7; 11.5) | 36 (24; 46) | 4.7 (3.8; 5.4) | 1 (-7; 8) | 3877† |
| Extended combined | 14.8 (13.5; 15.9) | 42 (30; 53) | 8.8 (7.7; 9.7) | 10 (5; 15) | 843 (525; 1556) |

TS = tongue swab, SoC = standard of care, USD = US dollars, ICER = incremental cost-effectiveness ratio, Ext. dominated = extendedly dominated.

* Sputum Xpert Ultra was extendedly dominated, meaning it was more costly and yielded fewer bacteriologically confirmed TB diagnoses than at least one combination of the other evaluated strategies. Strategies incorporating tongue swab-based testing using MiniDock MTB – either alone or in combination with sputum Xpert Ultra – achieved higher diagnostic yield at lower or comparable cost.

† The uncertainty interval for this ICER could not be calculated because incremental diagnoses were negative in some model iterations.

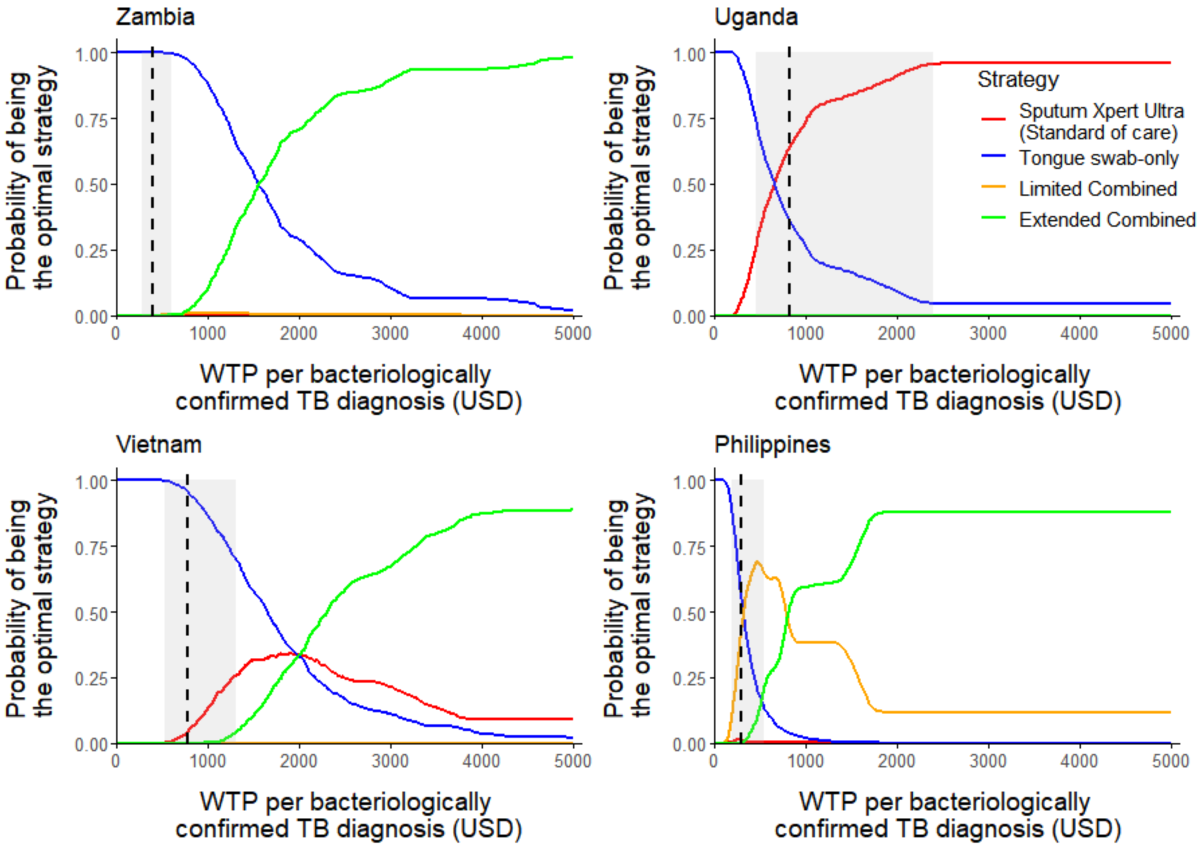


**Figure S1. Country-level cost-effectiveness acceptability curves for four tuberculosis (TB) diagnostic strategies.** Curves show the probability of each strategy being optimal across a range of willingness-to-pay (WTP) thresholds per additional bacteriologically confirmed TB diagnosis. The optimal strategy at each WTP is defined as the one with the highest expected net monetary benefit, based on probabilistic analysis.The analysis includes all four strategies: (1) *sputum Xpert Ultra* (standard of care), (2) *tongue swab-only* (MiniDock MTB), (3) *limited combined* strategy (sputum Xpert Ultra plus tongue swab MiniDock MTB for sputum-scarce individuals), and (4) *extended combined* strategy (sputum Xpert Ultra plus MiniDock MTB for sputum-scarce individuals and those with negative or indeterminate sputum results). In Uganda, tongue swab-based testing did not identify additional bacteriologically confirmed TB cases compared to sputum Xpert Ultra in the TSwaY study. However, the sample size was small (only 6 individuals tested positive on sputum Xpert Ultra, of whom 3 were also positive on tongue swab), limiting the ability to detect meaningful differences in diagnostic yield.

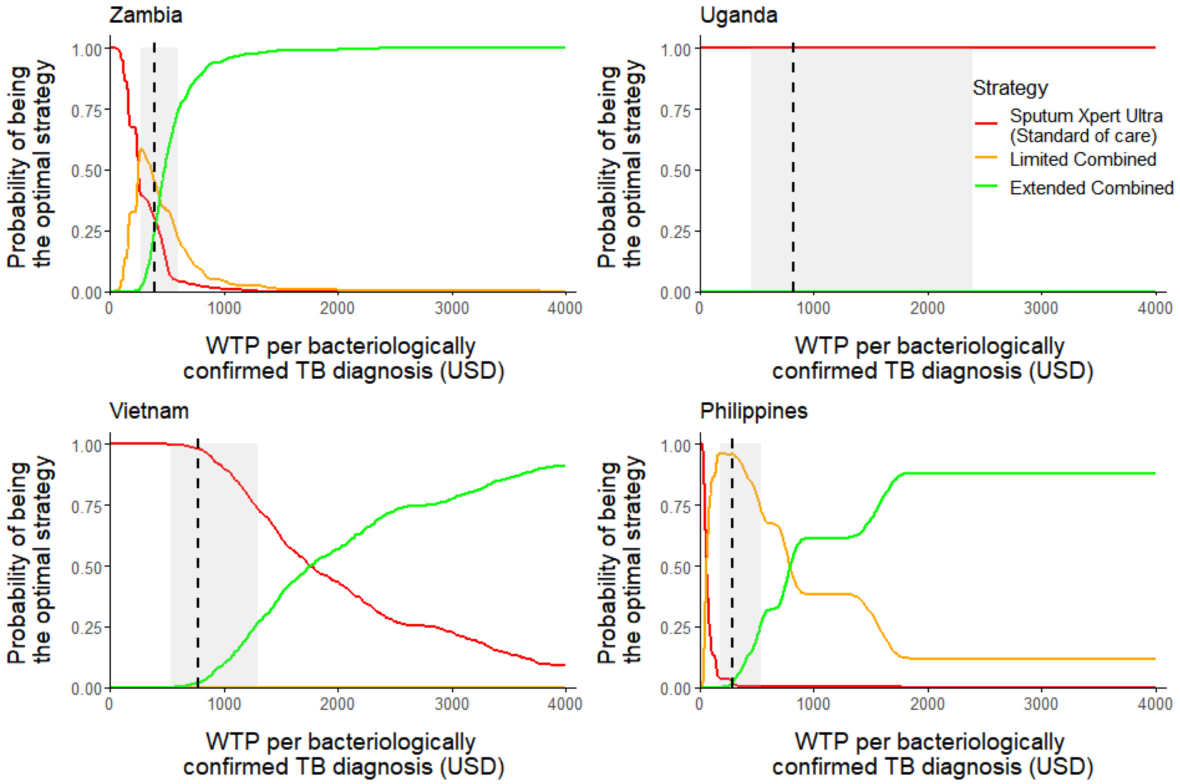


**Figure S2. Country-level cost-effectiveness acceptability curves for the standard of care (*sputum Xpert Ultra*) and the two combined diagnostic strategies (*limited combined*, *extended combined*), excluding the *tongue swab-only* strategy.**

Curves show the probability of each strategy being optimal across a range of willingness-to-pay (WTP) thresholds per additional bacteriologically confirmed TB diagnosis. The optimal strategy at each WTP is defined as the one with the highest expected net monetary benefit, based on probabilistic analysis. The analysis includes three strategies: (1) *sputum Xpert Ultra* (standard of care), (2) *limited combined* strategy (sputum Xpert Ultra plus tongue swab MiniDock MTB for sputum-scarce individuals), and (3) *extended combined* strategy (sputum Xpert Ultra plus MiniDock MTB for sputum-scarce individuals and those with negative or indeterminate sputum results). In Uganda, tongue swab-based testing did not identify additional bacteriologically confirmed TB cases compared to sputum Xpert Ultra in the TSwaY study. However, the sample size was small (only 6 individuals tested positive on sputum Xpert Ultra, of whom 3 were also positive on tongue swab), limiting the ability to detect meaningful differences in diagnostic yield.
